# Home and work-related injuries in Makwanpur district, Nepal: a household survey

**DOI:** 10.1101/2020.09.02.20186825

**Authors:** Puspa Raj Pant, Toity Deave, Amrit Banstola, Sumiksha Bhatta, Elisha Joshi, Dhruba Adhikari, Sunil Manandhar, Sunil Kumar Joshi, Julie Mytton

## Abstract

**Objective:** This paper describes the epidemiology of home- and work-related injuries, their mechanisms, inequalities and costs associated with these injuries.

**Methods:** A household survey was undertaken in three palikas (administrative areas) of Makwanpur district between April-June, 2019. Data were collected electronically on non-fatal injuries that occurred in the previous three months and fatal injuries that occurred in the previous five years.

**Findings:** 17,593 individuals were surveyed from 3,327 households. The injury rate was 8.0/1000 population for home injuries and 6.4/1000 for work-related injuries; 61.0% of home injuries were among females and 69.9% of work-related injuries among males. Forty-eight percent of all home injuries were due to falls (28 males (50.9%) and 40 females (46.5%)); burn/scalds were higher among females 15 (10.5%). Cuts and piercings accounted for 40% of all work-related injuries and 36.3% were falls. Injury incidence varied by ethnic group: home injuries were highest in Brahmin (12/1000) and work-related injuries highest in Rai groups (21.0 per 1000). The total mean cost of work-related injury was US$143.3 (SD 276.7), higher than for home injuries (US$130.4, SD 347.6). The number of home (n = 74, 64.9%) and work-related (n = 67, 77.9%) injuries were higher in families below the poverty line (<US$1.9) than families in the next income bracket (US$1.9–3.2)(home: n = 22, 19.3%; work: n = 11, 12.8%).

**Conclusions:** Home and occupational fall injuries are common. The inequalities in injury identified in our study by rurality, age, sex, income level and ethnic group can help target injury prevention interventions for vulnerable groups.

**What is already known on the subject:**

- Limited epidemiological data are available in Nepal about the location of injury, activity taking place when injury occurred and the costs of injury
- There is limited information about the distribution of injuries by ethnicity and income groups.

**What this study adds:**

- People from rural areas experience more injuries than people living in urban or city areas
- Manual workers sustain more work-related injuries than those in other occupations
- Certain ethnic groups have higher rates of injuries than other ethnic groups
- People with lower income experience more injuries than people with higher incomes
- Injury costs are high in relation to average per capita income.

## INTRODUCTION

Injury is relatively new on the public health agenda for Nepal. Hospital-based reports provide some information on injury-related mortality and morbidity, but community-based information derived from nationally representative samples is limited. Analysis of nationally representative census (2001) data reported an injury mortality rate of 30 per 100,000 population^1^ whereas the corresponding estimate for Nepal, produced by the Global Burden of Disease (GBD) study in 2017 was 59 per 100,000 population.^2^ In the absence of a robust death registration system in Nepal to record underlying causes of death, modelled data (such as those from the GBD study) are likely to provide the most robust estimates of injury deaths. Nepal’s Health Management Information System is the only routinely collected source of information on non-fatal injuries; this recorded 1.1 million outpatient department visits for injuries in the year 2017/18.^3^ The GBD study estimated 1.5 million injury outpatient visits in 2017, an annual rate of 50 per 1000 population.^2^

Household surveys conducted in different parts of Nepal at different times have found different estimates for non-fatal injuries; for example, in urban Dharan, minor injuries occurred in 35 per 1000 population (one month recall) and major injuries in 7 per 1000 population (one year recall).^4^ In rural Bhaktapur, the general injury rate was 29 per 1000.^5^ Minor injuries were defined as those resulting in a loss of less than 30 days of usual activity and major injuries as those resulting in a loss of 30 or more days. One study, conducted in rural eastern Nepal, reported an incidence of minor non-fatal injuries of 31 per 1000 population^6^; a nationwide survey, which used a lifetime experience of an injury, reported a high incidence of 131 per 1000 population non-fatal injuries.^7^ In Makwanpur, the incidence rate for injuries among children 0–17 years was 25 per 1000.^8^ The incidence of non-fatal home injuries among children aged less than 5 years living in rural areas of Makwanpur district was found to be as high as 232 per 1000.^9^ These sources identify the place of injury but not whether they were home- or work-related, nor have they reported on costs associated with injuries. A recent systematic review of injury research in Nepal identified an absence of evidence about inequalities in injury occurrence.^10^

This paper describes a study to explore the epidemiology of injuries related to home and work activities, inequalities in injury incidence and costs associated with injuries.

## METHODS

This study used a cross-sectional, community-based survey where data on injuries and the reported impact of these injuries on participants were collected face-to-face by trained data collectors between April and June, 2019. The setting was the Makwanpur district of Nepal, which includes three distinct administrative areas (known as ‘palika’) that has a topography representative of the country; research conducted there has the potential to be applicable to other districts.

A sample size of 3,325 was calculated using the standard formula suggested in guidelines provided by the United Nations for conducting community-based household surveys in low-income countries: a multi-stage, cluster sampling methodology was applied to selected households as the survey unit.^11^ The three palikas were selected purposively: one rural municipality, one municipality and one sub-metropolitan city. In each palika, four non-adjacent wards were selected purposively. The selection of households in the wards was proportional to the total number of households in each palika, systematic random sampling was used. All identified, eligible households were invited to take part. If there was no reply from a household at initial contact, the household was visited again at a different time of day. If unsuccessful at the second attempt, the next household on the list was selected.

For each household there was one respondent; the inclusion criteria for this person was that they were 18 years or over and, if a child was reported to have been injured, they were the main care-giver. Inclusion criteria for those reported to have been injured was that they could be any age but were, ordinarily, resident in one of the three palikas. Data were collected electronically on handheld tablet computers using REDCap data capture software by trained data collectors. The data collected included information about the household, about non-fatal injuries that had occurred in the previous three months and fatal injuries that had occurred in the previous five years, including type, location, circumstances and consequences of the injury sustained. Data collection tools were adapted from WHO guidelines on conducting community surveys on injuries and violence.^12^

An ‘injury’ was defined as an incident that resulted in a loss of at least one day of usual activity (e.g., absence from school) or one that required medical attention. A ‘home injury’ was defined as any injury that occurred within and around the home, not related to paid work or trade. ‘Work-related injury’ was defined as any injury that occurred whilst working in a paid job or for family subsistence, whether that work occurred at home, during a journey related to work or at a workplace, such as an office or factory. To collect data about ethnic groups, households were offered over 100 different ethnic groups to choose from. When describing ethnicity, we report the risks for the 6 most commonly reported ethnic groups and ‘other’.

The data were cleaned, coded and analysed using IBM SPSS Statistics 26. Descriptive statistics, percentages, rates and costs associated with injuries were calculated. For normally distributed data, the mean and standard deviation are presented, the median and the interquartile range (IQR) for skewed data. Differences between groups were investigated using non-parametric tests (chi-square). To ensure statistical disclosure control, cell numbers of less than 5 have been removed and, where necessary, eg., table 2, they are described in percentages. We used Nepal Census population size for ethnicities in the Makwanpur district to obtain denominator population for calculating injury incidence rates. Out-of-pocket (OOP) expenditure was the payment made by the household from its primary income or savings to cover the costs of injury (i.e., treatment costs and transportation costs). If payments were made by other methods (e.g., loan), they were deducted from the total costs of injury in order to calculate OOP expenditure.

Ethical approval for conducting this survey was obtained from the Nepal Health Research Council (Reg. no. 798/2018) and the University of the West of England Bristol Faculty Research Ethics Committee (reference no. HAS. 19.02.133).

## RESULTS

A total of 3,327 households (HHs) were surveyed within the three palikas, Bakaiya (499 HHs; 15.0%), Hetauda (2,256 HHs; 67.8%) and Thaha (572 HHs; 17.2%), including 17,593 individuals (49.5% females) (table 1). Seven households declined to participate in the survey. Within the previous three months, 358 (10.8%) households reported that someone had been injured. Bakaiya reported the highest proportion (n = 107, 21.4%) of survey households with an injured person. In contrast, the percentage of injured householders in Hetauda and Thaha was 8.3% (n = 187) and 11.2% (n = 64), respectively.

**Table 1:** Rates of home and work-related injuries in the survey population

| Category (whole sample- injured and uninjured) | Home injuries<br>n (rate/1000) | Work injuries<br>n (rate/1000) |
| --- | --- | --- |
| **Gender** |  |  |
| Female (n = 8,704) | 86 (9.9) | 34 (3.9) |
| Male (n = 8,889) | 55 (6.2) | 79 (8.9) |
| | | $\chi^2=0.513$, $p>0.05$ |
| **Palika** |  |  |
| Bakaiya (n= 3,023) | 30 (9.9) | 48 (15.9) |
| Hetauda (n= 11,485) | 90 (7.8) | 37 (3.2) |
| Thaha (n= 3,085) | 21 (6.8) | 28 (9.1) |
| | | $\chi^2=37.112$, $p<0.001$ |
| **Age groups** |  |  |
| Infants (n = 258) | * | * |
| 1-14 years (n = 3,738) | 47 (12.6) | 7 (1.9) |
| 15-24 years (n = 3,662) | 10 (2.7) | 21 (5.7) |
| 25-44 years (n = 5,617) | 36 (6.4) | 41 (7.3) |
| 45-64 years (n = 3,146) | 31 (9.9) | 36 (11.4) |
| 65+ years (n = 1,172) | 15 (12.8) | 8 (6.8) |
| **Family size** |  |  |
| Small family (up to 4 members) (n= 7,512) | 42 (5.6) | 35 (4.7) |
| Medium family (5-8 members) (n= 8,585) | 82 (9.6) | 65 (7.6) |
| Large family (9 or more members) (n= 1,496) | 17 (11.4) | 13 (8.7) |
| | | **$\chi^2=16.923, p<0.001$ |
| **Ethnicity\*** |  |  |
| Rai (n = 334) | * | 7 (21.0) |
| Brahmin (Hill) (n = 2,481) | 35 (14.1) | 22 (8.9) |
| Kami (n = 510) | * | * |
| Chhetri (n = 1,882) | 12 (6.4) | 16 (8.5) |
| Tamang (n = 8,409) | 55 (6.5) | 49 (5.8) |
| Newar (n = 1,091) | 11 (10.1) | * |
| All other ethnicities (n = 2,885) | 18 (6.2) | 16 (5.5) |
| **Total (N = 17,593)** | **141 (8.0)** | **113 (6.4)** |
| | | **$\chi^2=16.923, p<0.001$ |

Cells with less than 5 observations removed.
\* Estimated number of people for ethnicity based on the proportion of Census data 2011.
\*\* Post hoc analyses to test statistical difference in rates for both injury types combined.

Of the 394 reported injuries, 254 (64.5%) met the criteria for home or work-related injuries. There were 141 home-related injuries (8.0 per 1000 population) and 113 work-related injuries (6.4 per 1000 population). Sixty-one fatal injuries, due to any cause, were reported to have occurred over the previous 5 years, two of which were during the 3-month data collection period. Due to the low incidence, these cases are not reported further in this paper.

The age range of persons sustaining non-fatal injuries was between < 1 and 87 years with a median of 35 years (IQR 34)(table 1). The median age of injured males was 35 years (IQR 37) and 37.5 years (IQR 32) for females. Males had a slightly higher rate of injury (15.1 per 1000) compared to females (13.8 per 1000), however, home injuries were higher among females (9.9 per 1000) than males and work-injuries were higher among males (8.9 per 1000) compared to female. Within age groups, children aged 1–14 years and adults aged 45–64 years had the highest home injury rates (12.6 and 12.8 per 1000, respectively) whereas people aged 45–64 years had the highest work-related injuries (11.4 per 1000). Tamang were the largest ethnic group in the areas surveyed. Two ethnic groups, Rai (21.0 per 1000) and hill-living Brahmin (12.0 per 1000) reported the highest rates of work-related and home injuries, respectively) (table 1). Post hoc analyses found that there was a statistical difference in rates for both injury types between different ethnic groups (x^2^ = 16.923, p< 0.001) and between smaller and larger families (x^2^ = 25.175, p< 0.001) (table 1). Large families (9+ members) had higher rates of both home injuries (11.4 per 1000) and work-related injuries (8.7 per 1000 population) compared with small (1–4 members) and medium sized (5–8 members) families (table 1).

This study recorded 141 individuals (8.0 per 1000) with home-related injuries and 113 (6.4 per 1000) with work-related injuries. Injury incident rates for females were higher (9.9 per 1000) for home injuries compared to work-related injuries (3.9 per 1000); this was true in all three palikas. Work-related injury rates were higher among males (8.9 per 1000) compared to home injuries (6.2 per 1000). In rural Bakaiya, both the rate of home injuries (9.9 per 1000) and work-related injuries (15.9 per 1000) were higher than in Hetauda (urban) and Thaha (suburban)(table 1). Post hoc analyses found a statistical difference in rates for both injury types between different palikas (x^2^ = 37.112, p< 0.001).

Among all home injuries, 45.4% occurred in children (< 16 years) and the elderly (65+ years) whereas 86.7% of all work-related injuries were among people aged 15–64 years. Females reported more injuries at home than males at all ages except for children (< 16yrs) where boys had proportionally higher numbers of injuries than girls (n = 19, 40.4% and n = 28, 59.6%, respectively). For work-related injuries, males had, proportionally, a much higher number of injuries than females in all age groups (n = 79, 69.9% and n = 34, 30.1%, respectively), irrespective of the type of job (table 2).

**Table 2:** Inequalities in home and work-related injuries

|  | Home injuries |  |  | Work-related injuries |  |  |
| --- | --- | --- | --- | --- | --- | --- |
|  | Female<br>row % | Male<br>row % | Total<br>col % | Female<br>row % | Male<br>row % | Total<br>col % |
| **Palikas** |  |  |  |  |  |  |
| Bakaiya | 70.0% | 30.0% | 21.3% | 39.6% | 60.4% | 42.5% |
| Hetauda | 60.0% | 40.0% | 63.8% | 21.6% | 78.4% | 32.7% |
| Thaha | 52.4% | 47.6% | 14.9% | 25.0% | 75.0% | 24.8% |
| **Total** | **61.0%** | **39.0%** | **100%** | **30.1%** | **69.9%** | **100%** |
| **Age groups** |  |  |  |  |  |  |
| Infants | * | * | 1.4% | * | * | 0.0% |
| 1 to 14 years | 40.4% | 59.6% | 33.3% | * | 71.4% | 6.2% |
| 15 to 24 years | 80.0% | * | 7.1% | 42.9% | 57.1% | 18.6% |
| 25 to 44 years | 80.6% | 19.4% | 25.5% | 17.1% | 82.9% | 36.3% |
| 45 to 64 years | 67.7% | 32.3% | 22.0% | 36.1% | 63.9% | 31.9% |
| 65 years or over | 53.3% | 46.7% | 10.6% | * | 62.5% | 7.1% |
| **Total** | **61.0%** | **39.0%** | **100%** | **30.1%** | **69.9%** | **100%** |
| **Economic activity\*** |  |  |  |  |  |  |
| Mainly unemployed | 54.5% | 45.5% | 8.7% | * | 66.7% | 8.0% |
| Employed salaried | * | * | 4.7% | * | 87.5% | 14.3% |
| Home-maker | 92.1% | * | 29.9% | 60.0% | 40.0% | 22.3% |
| Agriculture | 42.9% | 57.1% | 11.0% | 35.0% | 65.0% | 17.9% |
| Student | 55.0% | 45.0% | 31.5% | 33.3% | 66.7% | 13.4% |
| Wage earners | * | * | 0.0% | * | 100.0% | 14.3% |
| All other occupation | 66.7% | 33.3% | 14.2% | * | 81.8% | 9.8% |
| **Total** | **66.1%** | **33.9%** | **100%** | **30.4%** | **69.6%** | **100%** |

| **Ethnicity** |  |  |  |  |  |  |
| --- | --- | --- | --- | --- | --- | --- |
| Tamang | 60.0% | 40.0% | 24.8% | 27.3% | 72.7% | 19.5% |
| Brahmin (Hill) | 66.7% | * | 8.5% | 31.3% | 68.8% | 14.2% |
| Newar | * | * | 4.3% | * | * | 0.0% |
| Chhetri | 72.7% | * | 7.8% | * | * | 2.7% |
| Kami | * | * | 2.8% | * | * | 6.2% |
| Rai | 60.0% | 40.0% | 39.0% | 32.7% | 67.3% | 43.4% |
| All other ethnicities | 55.6% | 44.4% | 12.8% | * | 87.5% | 14.2% |
| **Total** | **61.0%** | **39.0%** | **100%** | **30.1%** | **69.9%** | **100%** |
| **Mechanism of injury** |  |  |  |  |  |  |
| Road Traffic injury | * | * | 0.0% | * | 100.0% | 7.1% |
| Fall (including push or jump) | 58.8% | 41.2% | 48.2% | 31.7% | 68.3% | 36.3% |
| Poisoning | * | * | 2.1% | * | * | 0.0% |
| Animal/insect or reptiles' bite/sting | 75.0% | * | 8.5% | * | * | 2.7% |
| Fire, burn or scald | 75.0% | 25.0% | 14.2% | * | * | 1.8% |
| Electrocution | * | * | 0.7% | * | * | 1.8% |
| Cut, pierce, or impale | 47.8% | 52.2% | 16.3% | 42.2% | 57.8% | 39.8% |
| Injured by a blunt object | 72.7% | * | 7.8% | * | 91.7% | 10.6% |
| Other | * | * | 0.0% | * | 100.0% | 7.1% |
| **Total** | **58.8%** | **41.2%** | **48.2%** | **31.7%** | **68.3%** | **36.3%** |
| **Activity during injury<sup>¥</sup>** |  |  |  |  |  |  |
| Leisure or playing | 53.2% | 46.8% | 55.0% | * | * | 0.0% |
| Work | * | * | 0.0% | 31.3% | 68.8% | 85.0% |
| Other activities | 72.1% | 27.9% | 43.6% | * | * | 0.9% |
| Education | * | * | 0.7% | * | * | 0.0% |
| Travelling to and from work or school | * | * | 0.0% | * | 80.0% | 8.8% |
| Travelling for other purpose | * | * | 0.7% | * | * | 5.3% |
| **Total** | **61.4%** | **38.6%** | **100%** | **30.1%** | **69.9%** | **100%** |

Total usable sample size = 254 individuals. Cells with less than 5 observations removed.
\* information for 15 people is not available; <sup>¥</sup> one person declined to answer.

Table 2 also describes the economic activity, ethnicity, mechanism of injury and activity taking place when the injury occurred for the injured females and males. For those females who had home injuries (n = 86), 35 (40.7%) were home-makers and their most common home injuries were falls (n = 40,46.5%), burns or scalds (n = 15, 17.4%), cuts/piercings (n = ll, 12.8%) and animal-related (n = 9, 10.5%). For males who had a home injury (n = 55), their most common injuries were falls (n = 28, 50.9%) and cuts/piercings (n = 12, 21.8%).

### Costs of injury

The costs of injury and out of pocket (OOP) expenditure incurred by the 139 injured persons are presented in table 3. The total mean cost of work-related injury was US$ 143.3 (SD 276.7) higher than for home injuries (US$ 130.4, SD 347.6). The mean OOP expenditure was higher for home injuries than work-related injuries (US$ 83.1, SD 199.6 and US$ 75.6, 127.7, respectively) and accounted for 57.9% of the total costs of injury.

**Table 3.** Total costs and out-of-pocket payment (OOP) (in US$)

|  | Home-related (n = 86)<br>**Mean (SD)** | Work related (n =53)<br>**Mean (SD)** |
| --- | --- | --- |
| **Treatment costs** | 122.7 (342.8) | 128.4 (260.4) |
| **Transport costs** | 7.7 (21.6) | 14.9 (42.6) |
| **Total costs** | 130.4 (347.7) | 143.3 (276.8) |
| **OOP Expenditure** | 83.2 (199.8) | 75.6 (127.8) |

*SD, Standard Deviation; 1 US\$ = NRs 110 (June 2019)*

In our study, monthly income was known for 200 (out of those 254 with home or work-related injuries) and 141 (70.5%) had an income of below 1.9 USD per day (living below the World Bank poverty line^13^). In Thaha, all of the injured persons (n = 27) were in this category while 78.0% (n = 50) in Bakaiya and 61.0% (n = 123) in Hetauda were living below the poverty line. Among those with work-related injuries, 77.9% (n = 67) were earning less than 1.9USD daily while this proportion was lower (64.9%, n = 74) for those with home injuries. The results also suggest that both home and work-related injuries were higher among the low-income earners compared to those participants in the other two income brackets (p = 3.98; df 1; x^2^ = 0.046)(table 4).

**Table 4:**
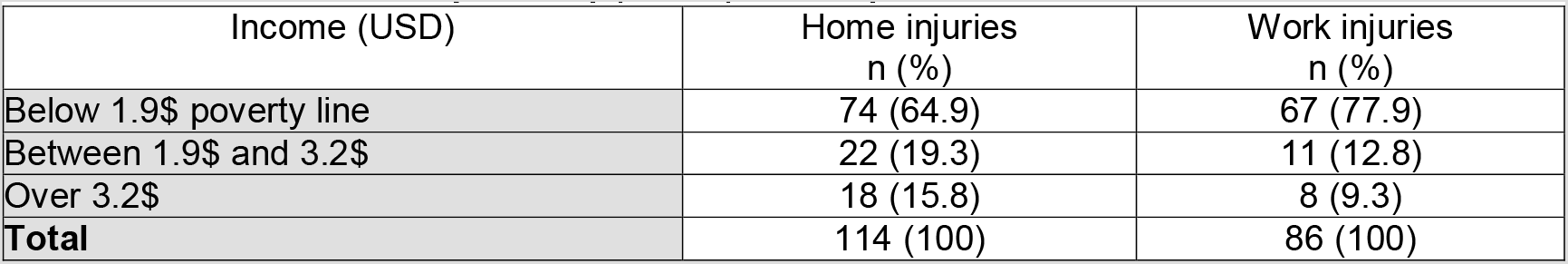
Distribution of injuries by per capita daily income

## DISCUSSION

To our knowledge, this is the first comprehensive study to identify, describe and study distribution and inequalities of injuries related to home and work in Nepal. The survey data were collected from three different palikas that were typical of the Makwanpur district of Nepal. The findings suggest that there are particular at-risk groups for both home and work-related injuries. Out of all the palikas, the rural area (Bakaiya) had the highest rates of injuries, which confirms findings from other South Asian countries where rural areas also had high rates of injury.^14^^15^ Household surveys previously conducted in different parts of Nepal recorded injury rates ranging between 7 per 1000 to 31 per 1000 per year.^4^–^7^ Whilst these studies recorded minor and major injuries, none of them differentiated between home or work-related injuries.

High rates of home injuries in children in Nepal have previously been reported^9^ and have also been found in Bangladesh.^15^ Our study found that falls and cuts/piercings were the main mechanism for both home- and work-related injuries. Fall injuries have also been the most commonly reported injuries in two studies in India^16^^17^ and Pakistan^18^ where the children under the age of 15 years were at high risk of fall injuries. However, falls were found to be an equally serious problem for the elderly population in Bangladesh.^19^ Unlike Bangladesh, in our study we found falls were common among children 1–14 years (33.8%) and just 14.7% were among older people aged over 65 years. In our study, non-fatal burns and scalds were more common home injuries (14.2%) than in Bangladesh (< 4%)^20^ but more similar to India (17%)^17^; they are a major cause of morbidity and disability.

In relation to home injuries, the percentage of all injuries that were home-related in our study are similar to those reported in a recent systematic review conducted in Nepal.^10^ Work-related injuries were much higher in our study (28.7%) than in those studies included in the review (11–19%).^10^ Earlier studies considered the place of injury occurrence but our study added context and applied operational definitions to the place of injury occurrence, e.g., home- or work-related. The high number of work-related injuries could be explained by two reasons. Firstly, the definition we used included unpaid work because it contributes to a family’s subsistence by providing an alternative to paid work for that family.^20^ For example, many families living in rural Nepal keep their own livestock and grow their own food. Secondly, as per the topographic and socio-demographic situation of the survey district, many people were engaged in manual and informal/unpaid work; the latter is common in rural areas. We also explored injuries in relation to economic activity, ethnicity and costs. In relation to agricultural injuries, our findings support those from an earlier Nepalese study where 69% of farmers reported an injury in the past 12 months.^21^

Our study found 6.2% of all work-related injuries were among children below 15 years. Although we categorised children sustained work-related injuries, as per our working definition, most of these injuries were associated with unpaid work whilst they tended the family livestock and undertook animal husbandry.

Nepal is a diverse country in respect of ethnicities with 125 ethnicities recorded in the last census.^22^ A previous study undertaken in the same district as our survey reported frequencies for emergency department (ED) attendance. They also reported that there was a higher proportion of *Tamang* and *Brahmin* ethnic groups who attended EDs with an injury.^23^ However, when rates of injury were calculated, we found that the *Brahmin* and *Rai* ethnicities had the highest rates of home and work-related injury, respectively. The previous study looked at ED attendance and reported frequencies whereas our study was a population survey and we report rates of injury. *Tamang* were the majority ethnic group in Makwanpur district and they had the highest absolute number of injuries^22^ but their rates of injury were lower than *Rai* and *Brahmin* groups. Most of the injured *Rai* ethnic group in our study live in the Bakaiya (rural) palika but this *Rai* community *(Danuwar or Dewas Rai)* is different to those who live in Eastern Nepal *(Kirant Rai)*.^24^ The inequality in injury incidence by ethnic group suggests that this should be investigated further.

The costs of injuries in our sample were high in relation to income and were higher for work-related injuries than home injuries. To the best of our knowledge, there have been no previous studies in South Asia that have explored costs of home and work-related injuries.^25^ The World Bank estimated the rate of poverty (defined as an income of $1.90 per person per day) to be 8 percent in Nepal in 2019.^13^ Our study we asked the respondents about the family monthly income, this was then calculated per capita (USD). We found that the majority of home and work-related injuries happened to those who were living below 1.9USD per day.

### Strengths and limitations

There are three principal strengths of this study, the first being the representative nature of the households that were recruited due to the sampling method used. Secondly, trained data collectors were committed and well supervised which led to complete data being collected with very few missing data. Thirdly, households were offered over 100 different ethnic groups to choose from, thus making this the first comprehensive household survey to explore injury by ethnic group in Nepal. Participants were willing to report their ethnic group and our findings indicate that there are inequalities by ethnic group that warrant further investigation. Although previous injury household surveys provided information on the location of injury, there are no studies that reported home- and work-related injuries. One limitation of our study was that the participants were asked to report non-fatal injuries that happened in the previous three months and five years for fatal injuries. Whilst this was thought to be feasible for participants, there is a chance of recall bias as people tend to forget more minor injuries^26^ A second limitation was the cross-sectional nature of this study so we cannot comment on causality. As in any observational study, the potential for unmeasured confounding exists and the nature of non-fatal injuries may have differed if it had been conducted at a different time of year. It is likely that there will be seasonal variation, for example, during the monsoon when travel is reduced and potentially more dangerous, there are monsoon-related injuries, such as drowning.

## CONCLUSION

Inequalities in injury were found in relation to rural living, age, sex, low income and specific ethnic groups. Falls and cuts/piercings were the main mechanisms of injury for both home-and work-related injuries; non-fatal burns and scalds were common home injuries. This study highlights the need for the development and implementation of injury prevention programmes and policies that target geographic and socio-demographic differences in injury incidence and particular home- and work-related injuries.

## Data Availability

Data is available upon request

## ACKNOWLEDGEMENTS

We acknowledge the support of all three Municipality authorities for approving to conduct this study. We are grateful to all the staff at Mother and Infant Research Activities and the data collectors for their support with recruitment of the study participants and logistics to conduct the data collection. We are thankful to all the participants of our study who consented to participate. We would like to acknowledge the support of the wider research team at NIHR Global Health Research Group on Nepal Injury Research UK and Nepal team who have supported the study.

## CONTRIBUTORS

PRP, TD, SKJ and JM contributed to the conception and design of the study. PRP and TD drafted the protocol design, methods and data analysis plan. EJ, SB, DA, PRP and SM supported the data collection. PRP and TD led the analysis, interpretation of data and drafted the article. PRP, TD and JM drafted and finalised the manuscript. All the authors contributed to drafts and approved the final manuscript.

## REFERENCES

1. Sharma G. Leading causes of mortality from diseases and injury in Nepal: a report from national census sample survey. Journal of institute of medicine 2006;28(1)

2. GBD Collaborative Network. Global Burden of Disease Study 2017 (GBD 2017) Results. Seattle, United States, 2018.

3. MoHP. HMIS Database 2074/75 by Local Government Kathmandu2020 [updated 25 March 2020. Available from: https://dohs.gov.np/ihims-raw-data/ accessed 26 March 2020.

4. Ghimire A, Nagesh S, Jha N, et al. An epidemiological study of injury among urban population. Kathmandu University Medical Journal 2009;7(28):402–07.

5. Aryal UR, Vaidya A, Shakya-Vaidya S, et al. Establishing a health demographic surveillance site in Bhaktapur district, Nepal: initial experiences and findings. BMC Research Notes2012;5:489. doi: https://dx.doi.org/10.1186/1756-0500-5-489

6. Khanal V, Upreti R, Oli U, et al. Prevalence of injury and its associated factors in a rural area of eastern Nepal. Journal of Chitwan Medical College 2017;6(4):7–13.

7. Gupta S, Wong EG, Nepal S, et al. Injury prevalence and causality in developing nations: Results from a countrywide population-based survey in Nepal. Surgery 2015;157(5):843–49. doi: https://dx.doi.Org/10.1016/i.surg.2014.12.020

8. Pant PR, Towner E, Ellis M, et al. Epidemiology of Unintentional Child Injuries in the Makwanpur District of Nepal: A Household Survey. International Journal of Environmental Research & Public Health 2015;12(12):15118–28. doi: https://dx.doi.org/10.3390/iierphl21214967

9. Bhatta S, Mytton J, Deave T. Environmental change interventions to prevent unintentional home injuries among children in Low-and Middle-Income Countries: a systematic review and meta-analysis. Child: care, health and development 2020

10. Mytton J, Bhatta S, Thorne M, et al. Understanding the burden of injuries in Nepal: A systematic review of published studies. Cogent Medicine 2019;6(1):1673654.

11. United Nations. Designing Household Survey Samples: Practical Guidelines. New York: Department of Economic and Social Affairs, Statistics Division 2008.

12. World Health 0. Guidelines for conducting community surveys on injuries and violence / edited by D. Sethi… [et al.]. Geneva: World Health Organization, 2004.

13. World Bank. The World Bank in Nepal Kathmandu: The World Bank; 2020 [updated 12 April 2020. Available from: https://www.worldbank.org/en/countrv/nepal/overview accessed July 20 2020.

14. Jagnoor J, Suraweera W, Keay L, et al. Unintentional injury mortality in India, 2005: nationally representative mortality survey of 1.1 million homes. BMC public health 2012;12(1):487.

15. Giashuddin SM, Rahman A, Rahman F, et al. Socioeconomic inequality in child injury in Bangladesh-implication for developing countries. International journal for equity in health 2009;8(1):7.

16. Cardona Ivi, Joshi R, Ivers RQ, et al. The burden of fatal and non-fatal injury in rural India. Injury prevention 2008;14(4):232–37.

17. Bhanderi DJ, Choudhary S. A study of occurrence of domestic accidents in semi-urban community. Indian journal of community medicine: official publication of Indian Association of Preventive & Social Medicine 2008;33(2):104.

18. Bachani AM, Ghaffar A, Hyder AA. Burden of fall injuries in Pakistan—analysis of the National Injury Survey of Pakistan. Eastern Mediterranean health journal 2011;17(5)

19. Wadhwaniya S, Alonge O, Ul Baset M, et al. Epidemiology of fall injury in rural Bangladesh. International journal of environmental research and public health2017;14(8):900.

20. Swiebel J. Unpaid work and policy-making: Towards a broader perspective of work and employment (A discussion paper of the United Nations Department of Economic and Social Affairs). DESA Discussion Paper. New York: United Nations, 1999.

21. Bhattarai D, Singh SB, Baral D, et al. Work-related injuries among farmers: a cross-sectional study from rural Nepal. Journal of occupational medicine and toxicology 2016;11(1):48.

22. CBS. Population monograph of Nepal Vol II: Social Demography. Kathmandu: Central Bureau of Statistics, 2014.

23. Bhatta S, Pant PR, Mytton J. Usefulness of hospital emergency department records to explore access to injury care in Nepal. International journal of emergency medicine 2016;9(1):21.

24. Shackelford S. A sociolinguistic study of Dewas Rai and Danuwar. SIL Electronic Survey Report.Dallas. Texas: SIL International, 2019:162.

25. Zaloshnja E, Miller TR, Lawrence BA, et al. The costs of unintentional home injuries. American journal of preventive medicine 2005;28(1):88–94.

26. Moshiro C, Heuch I, Astrøm AN, et al. Effect of recall on estimation of non-fatal injury rates: a community based study in Tanzania. Injury Prevention 2005;11 (1):48–52. doi: 10.1136/ip.2004.005645

